# Exploring novel inflammation-related genetic and hematological predictors of response to neoadjuvant chemoradiotherapy in locally advanced rectal cancer

**DOI:** 10.1101/2023.06.20.23291673

**Authors:** Mladen Marinkovic, Suzana Stojanovic-Rundic, Aleksandra Stanojevic, Marija Ostojic, Dusica Gavrilovic, Radmila Jankovic, Natasa Maksimovic, Rafael Stroggilos, Jerome Zoidakis, Sergi Castellví-Bel, Remond J.A. Fijneman, Milena Cavic

## Abstract

**Background:** The standard initial treatment for locally advanced rectal cancer (LARC) is neoadjuvant chemoradiotherapy (nCRT). In order to select patients who would benefit the most from nCRT, there is a strong need for predictive biomarkers. The aim of this study was to evaluate the role of clinical, pathological, radiological, inflammation-related genetic, and hematological parameters in the prediction of response after nCRT.

**Methods:** *In silico* analysis of published transcriptomics datasets was conducted to identify the best candidate genes, whose expression will be measured using quantitative Real Time PCR (qRT-PCR) in pretreatment formaline-fixed paraffin-embedded (FFPE) samples. In this study, 75 patients with LARC, between June 2020 and January 2022, were prospectively included. Patients were assessed for tumor response in the 8th week after nCRT completion with pelvic MRI scan and rigid proctoscopy. For patients with a clinical complete response (cCR) and initially distant located tumor no immediate surgery was suggested (“watch and wait” approach). The response after surgery was assessed using histopathological tumor regression grading (TRG) categories from postoperative specimens by Mandard. Responders (R) were defined as patients with cCR without operative treatment, and those with TRG 1 and TRG 2 postoperative categories. Non-responders (NR) were patients classified as TRG 3-5.

**Results:** Responders group comprised 35 patients (46.6%) and NR group included 53.4% of patients. Analysis of published transcriptomics data identified genes that could predict response to treatment and their significance was assessed in our cohort by qRT-PCR. When comparison was made in the subgroup of patients who were operated (TRG1 vs. TRG4), the expression of IDO1 was significantly deregulated (*p*<0.05). Among hematological parameters between R and NR a significant difference in the response was detected for neutrophil-to-monocyte ratio (NMR), initial basophil, eosinophil and monocyte counts (*p*<0.01). According to MRI findings, non-responders were more often presented with extramural vascular invasion (*p*<0.05).

**Conclusion:** Based on logistic regression model, factors associated with favorable response to nCRT were found to be tumor morphology as well as hematological parameters which can be easily and routinely derived from initial laboratory results (NMR, eosinophil, basophil and monocyte counts) in a minimally invasive manner. Using various metrics, an aggregated score of the initial eosinophil, basophil, and monocyte counts demonstrated the best predictive performance.

## Introduction

In 2020, colorectal cancer (CRC) was the third most common malignant disease with 1.9 million new cases worldwide ^1^. With 0.9 million deaths, it held the second place of cancer-related mortality causes in 2020 ^1^. In Serbia, in 2020, there were 2,956 new cases and a total of 1,493 deaths related to rectal cancer, which placed Serbia in the group of countries with a high incidence and mortality rate for this disease ^1^. In the majority of cases, it is diagnosed in advanced stages, when treatment options are limited. In this regard, in the past we have profiled the diagnostic, prognostic and predictive factors for cancers of the digestive system, leading to improved research strategies for patient management and care ^2–7^. However, there is a need for better primary prevention, more effective screening program, diagnosis at an earlier stage of the disease and improvement of existing treatment modalities in our country and on a global level.

The standard treatment for locally advanced rectal cancer (LARC) is neoadjuvant chemoradiotherapy (nCRT) followed by total mesorectal excision with or without adjuvant chemotherapy. The pathologic complete response (pCR) after nCRT is achieved in 10-30% of cases^8^. It has been reported that pCR, independent of the initial clinical T and N stage of the disease, was associated with better local and distant disease control, as well as longer disease-free and overall survival ^9^. Other reports showed that radical surgical treatment was related to significant morbidity, including postoperative complications ^10^. Further investigations were directed towards less invasive surgical treatment or avoiding surgery (“watch and wait” approach) in patients with favorable response to nCRT, in order to improve the quality of life. Since 2004, a group of researchers led by Angelita Habr-Gama have contributed greatly in this area by pointing out the effectiveness and safety of this approach ^11^. The current management of LARC uses the clinical complete response (cCR) as the point of reference for identifying patients for whom a non-operative approach may be a viable option ^12^. However, the clinical response poorly correlates with the pathologic response ^13^.

Other research trends in this field were dedicated to prolonging the period between completion of neoadjuvant treatment and surgery, changing the type and regimen of chemotherapy, as well as increasing the radiotherapy doses. These approaches aimed to achieve a higher percentage of good response to the initial treatment. As not all patients will benefit from these treatment modifications, there is a need to categorize them initially before treatment. In order to select patients who would benefit the most from a neoadjuvant treatment, there is a strong demand to discover and characterize predictive biomarkers. Despite numerous studies in this field, until now no molecular marker has been implemented as a diagnostic or predictive parameter in routine clinical practice of LARC. This is stressed by the fact that there was not enough matching regarding results of published studies in this area and only two genes (MMP4 and FLNA) were shown to be significant in more than one study ^14^. Limitations of previous studies included a small number of patients, the absence of reproducibility of measurements, the use of different methodologies, the retrospective nature of the studies, the heterogeneity of the studied groups and applied treatment modalities, as well as the lack of verification of the findings. Further research was aimed at examining the cumulative effect of molecular markers in combination with radiological and clinical data. An example of such successful research is the examination of the correlation between the expression of three protein molecular markers (c-MYC, PCNA and TIMP1) and magnetic resonance imaging (MRI) parameters ^15^.

The association between inflammatory bowel disease and the higher risk of developing colorectal cancer is well known ^16, 17^. Also, there is evidence of the role of inflammation in sporadic colorectal cancer ^18, 19^. Chronic inflammation in the tumor microenvironment has also been shown to favorize tumor growth and invasiveness and stimulate synthesis of epithelial to mesenchymal transition promoting transcription factors ^20^. Yet, no inflammation-related genetic or circulating biomarkers have been investigated in detail or established as predictive parameters in the LARC setting so far.

The aim of this study was to evaluate the role of clinical, pathological, radiological, inflammation-related genetic and hematological parameters in prediction of response after nCRT in patients with LARC.

## Methods

### *In silico* analysis of published transcriptomics datasets

*In silico* analysis of published transcriptomics datasets was conducted to identify the best candidate genes, whose expression will be measured using quantitative Real Time PCR (qRT-PCR) in pretreatment formaline-fixed paraffin-embedded (FFPE) samples.

Gene expression patterns were analyzed using publicly available datasets. By searching the public database the National Center for Biotechnology Information Gene Expression Omnibus (NCBI GEO) using key words rectal cancer, chemoradiotherapy and response to treatment, five adequate sets of data that analyzed pretreatment samples were identified: GSE45404, GSE68204, GSE139255, GSE46862 and GSE3493 ^21–25^. Three datasets were selected where inflammatory response significantly correlated with treatment outcome to nCRT. Gene expression profiles of GSE46862, GSE139255, and GSE45404_570 were obtained from GEO database. The total number of patients of each dataset was 69, 156, and 42 respectively. In all selected studies, the response to treatment was classified according to pathohistological tumor regression grading (TRG) categories from the postoperative specimen using Mandard scoring system. Patients were subdivided into responders (TRG 1-2) and non-responders (TRG 3-5). Microarray data were processed and normalized with the Robust Multichip Average method ^26^. Analysis for statistically significant differences between the two groups was conducted using the standard moderated t-test from the limma package ^27, 28^. Gene set enrichment analysis (GSEA) was performed on selected datasets, and Hallmark, Kyoto encyclopedia of genes and genomes (KEGG), and Reactome gene sets were used to identify pathway alterations in patients who responded well to the therapy (TRG 1-2) versus those who did not (TRG 3-5) ^29, 30^.

Next, the top 100 genes from selected datasets, ranked by the default Signal2Noise metric used in previously described GSEA analysis, were extracted and overlapped using Venn diagram software. Cytoscape (version 3.10.0) was applied as bioinformatics software to evaluate the potential correlation between finally selected genes ^31, 32^.

### Patient characteristics, treatment and follow-up

In this study 75 patients with LARC treated at the Institute for Oncology and Radiology of Serbia, between June 2020 and January 2022, were prospectively included. The inclusion criteria were histopathologically verified adenocarcinoma of the rectum, with a distant margin up to 12 cm from the anal verge by rigid proctoscopy. LARC was defined as T3-T4N0 or any T stage N positive. Pretreatment evaluation included an abdominal and pelvic MRI scan and a computed tomography (CT) scan or X ray of the chest. All patients were treated with long-course nCRT. Radiotherapy (RT) was delivered using volumetric modulated arc therapy-simultaneous integrated boost technique (VMAT-SIB). The dose to mesorectum and pelvic lymph nodes was 45 Gy (1.8 Gy/fraction). A SIB was delivered on macroscopic disease region expanded with 2 cm margin with a total dose of 54 Gy (2.16 Gy/fraction). Concomitant chemotherapy started on the first day of RT and was administered during the first and the fifth week of RT. The chemotherapy regimen included: 5-FU (350 mg/m2 on the first day of the first and fifth week of RT) and Leucovorine (25 mg/m2 daily, 5 days of the first and fifth week of RT).

Patients were assessed for tumor response in the 8th week after nCRT completion with pelvic MRI scan, rigid proctoscopy and digital rectal examination. For patients with cCR and initially distant located tumor no immediate radical surgery was suggested and they were enrolled in a strict follow-up program (“watch and wait” approach). Patients with cCR where sphincter preservation surgery treatment can be delivered, were referred to surgical resection between weeks 8 and 12 from nCRT completion. For patients with partial response (PR), surgery was delayed until week 12-15, approximately. The pathohistological response after surgery was assessed according to classification by Mandard. The response to treatment was classified according to pathohistological TRG categories from the postoperative specimen. Responders were defined as patients with cCR without operative treatment, and those with TRG 1 and TRG 2 postoperative categories. Non-responders were patients classified as TRG 3-5.

Formalin-fixed paraffin-embedded (FFPE) samples taken at the time of disease diagnosis were collected. The project was approved by the Ethics Committee of the Institute for Oncology and Radiology of Serbia (Approval No. 2211-01 from 11.06.2020.) and Ethics Committee of the Faculty of Medicine, University of Belgrade (Approval No. 1322/XII-17 from 03.12.2020.). All patients signed an informed consent.

Before initiation of treatment, ethylenediaminetetraacetic acid (EDTA) peripheral blood was drawn by venipuncture and hematological parameters were derived from the absolute differential counts of a complete blood count (CBC). The neutrophil-to-lymphocyte ratio (NLR) was calculated as a ratio of circulating neutrophil and lymphocyte counts, and the platelet-to-lymphocyte ratio (PLR) was defined as the absolute count of platelets divided by the absolute lymphocyte count. The derived neutrophil-to-lymphocyte ratio (dNLR) was calculated as absolute neutrophil count divided by absolute leukocyte minus absolute neutrophil count. The lymphocyte-to-monocyte ratio (LMR), platelet-to-monocyte ratio (PMR), and neutrophil-to-monocyte ratio (NMR) were also analyzed. Patients’ pre-treatment hemoglobin levels were obtained. The staging of the tumor was assessed according to the eighth edition of the Union for International Cancer Control (UICC) TNM staging system for rectal cancer ^33^. The general condition of the patients was classified using the Eastern cooperative oncology group (ECOG) Scale of Performance Status ^34^.

### RNA isolation and cDNA synthesis

Total RNA was isolated from 2-5 10 μm thick FFPE tissue sections using RNeasy FFPE Kit (Qiagen, Manchester, UK). RNA quality and concentration were determined spectrophotometrically using BioSpec-nano (Shimadzu Scientific Instruments, Kyoto. Japan). The complementary DNA (cDNA) was accessed from 1 µg total RNA using random primers and MultiScribeTM Reverse Transcriptase (50 U/µL) from the High-Capacity cDNA Reverse Transcription kit (Applied Biosystems, Foster City, CA, USA). The reaction was performed in 20 µL, using the following program: 25°C for 10 min, 37°C for 120 min, and inactivation at 85°C for 5 min.

### Quantitative Real Time PCR (qRT-PCR)

The mRNA levels of IL6 (RefSeq. NM_000600.5), CXCL9 (RefSeq. NM_002416.3), IDO1 (RefSeq. NM_002164.6) and CYBB (RefSeq. NM_000397.4) were detected by quantitative real-time PCR (qRT-PCR) using oligonucleotides primers (Integrated DNA Technology, Coralville, Iowa, USA) previously designed using NCBI Primer Blast and SybrGreen Gene Expression Master Mix (Applied Biosystems), on ABI Prism 7500 Sequence Detection System (Applied Biosystems). The thermal cycling conditions consisted of an UDG activation at 50°C, initial denaturation step at 95°C for 2 min followed by 40 cycles of denaturation (15 sec at 95°C) and annealing/extension (1 min at 60°C). All experiments were performed in duplicate, including non-template controls in each amplification. Gene expression data were normalized to glyceraldehyde-3-phosphate dehydrogenase (GAPDH, RefSeq. NM_002046.5). Data was analyzed using the classical delta-delta-Ct method, and results expressed in relative units.

### Statistical analysis

For normal distribution data testing, the Kolmogorov-Smirnov and Shapiro-Wilk tests were used. Descriptive methods (frequencies, percentage, mean, median, standard deviation (SD) and range) were used to summarize the data. The statistical significance level was set at *p*<0.05. For comparison of disease and treatment characteristics among different subgroups the Wilcoxon rank sum, Pearson chi-square and Fisher exact tests were used. Also, for evaluating potential predictors of the response, univariate and multivariate logistic regression was used (odds ratio with 95% CI for description, Likelihood Ratio and Wild test), and the responders versus non-responders was set as a dependent variable. We evaluated the sensitivity, specificity, positive predictive value, negative predictive value, and predictive accuracy for clinical assessment of disease presence in comparison with pathohistological response as a gold standard in group of patients where operative treatment was conducted ^35^. The Receiver Operating Characteristics curve (ROC) methods were applied to investigate the discriminative potential of NLR, PLR, dNLR, LMR, PMR, NMR, initial basophil, eosinophil and monocyte counts, for the good response to treatment (AUC ROC-Area Under the ROC curve according DeLong’s method; Likelihood ratio test for AUC ROC; the best cut-off value was set as value with maximum sensitivity and specificity). The statistical analysis was performed using the program R (version 3.3.2 (2016-10-31) --“Sincere Pumpkin Patch”; Copyright (C) 2016 The R Foundation for Statistical Computing; Platform: x86_64-w64-mingw32/× 64 (64-bit); downloaded: January 21, 2021). In the search for a measure that outperforms the individual variables, numerical variables that remained significant in the multivariate analysis were utilized to create the composite scores. The predictive power of these scores was then tested using the AUC, the Area Under Precision-Recall Curve (AUCRP), the Root Mean Square Error (RMSE) as a metric (using the ROCR package) and a random forest classifier (using the randomForest package, with the MeanDecreaseAccuracy metric) ^36^.

## Results

After extensive search of the GEO database according to the key words rectal cancer, chemoradiotherapy and response to treatment, three gene expression datasets were finally obtained. Volcano plots were employed to identify genes that exhibited statistically significant differential expression between responders and non-responders, as determined by the adjusted p-value, among selected datasets (Figure 1). Significant alteration defined as those with adjusted p-value lower then 0.05 was obtained only for GSE 139255 dataset. The results of the differential expression analysis within GSE 139255 are reported in Supplementary Material 1. While KEGG and Reactome GSEA analysis yielded no overlap among the selected datasets, the Hallmark analysis exhibited consistent and significant parameters across two datasets. Results of GSEA Hallmark analysis presented in Table 1 showed parameters which reached significant levels within GSE46862 and GSE139255 datasets in relation to Hallmark inflammatory response pathway (NOM p-value < 0.05, FDR q-value < 0.25). Enrichment plots were used to present the expression of genes in selected datasets in Figure 2.

**Figure 1.**
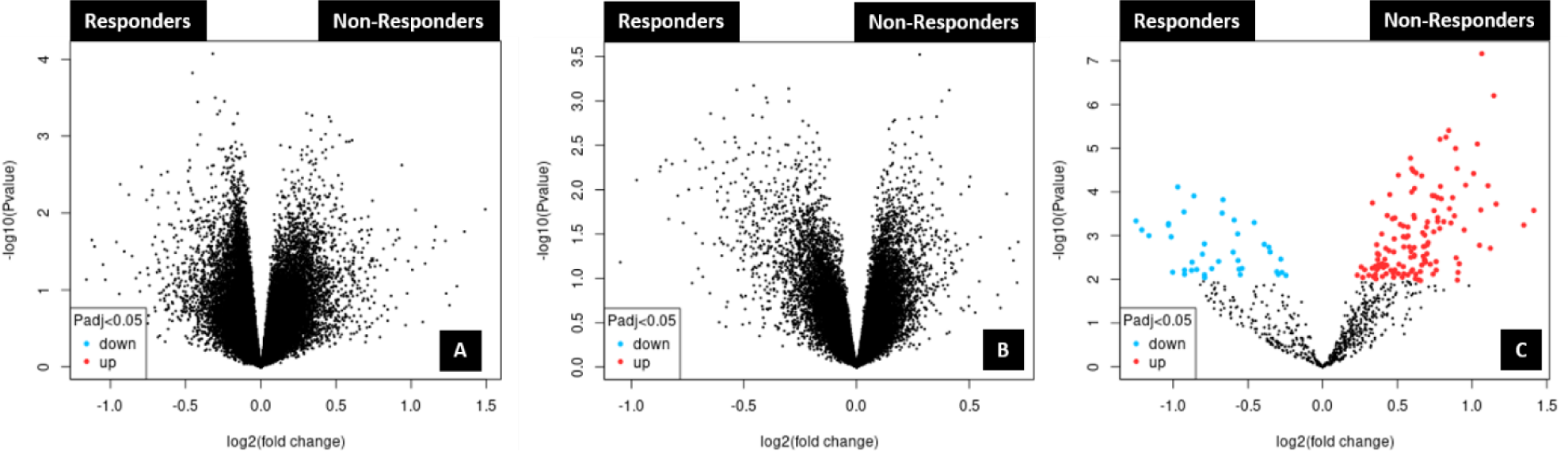
**Volcano plots for selected datasets: GSE45404_570 (A); GSE46862 (B); GSE139255 (C).**

**Figure 2.**
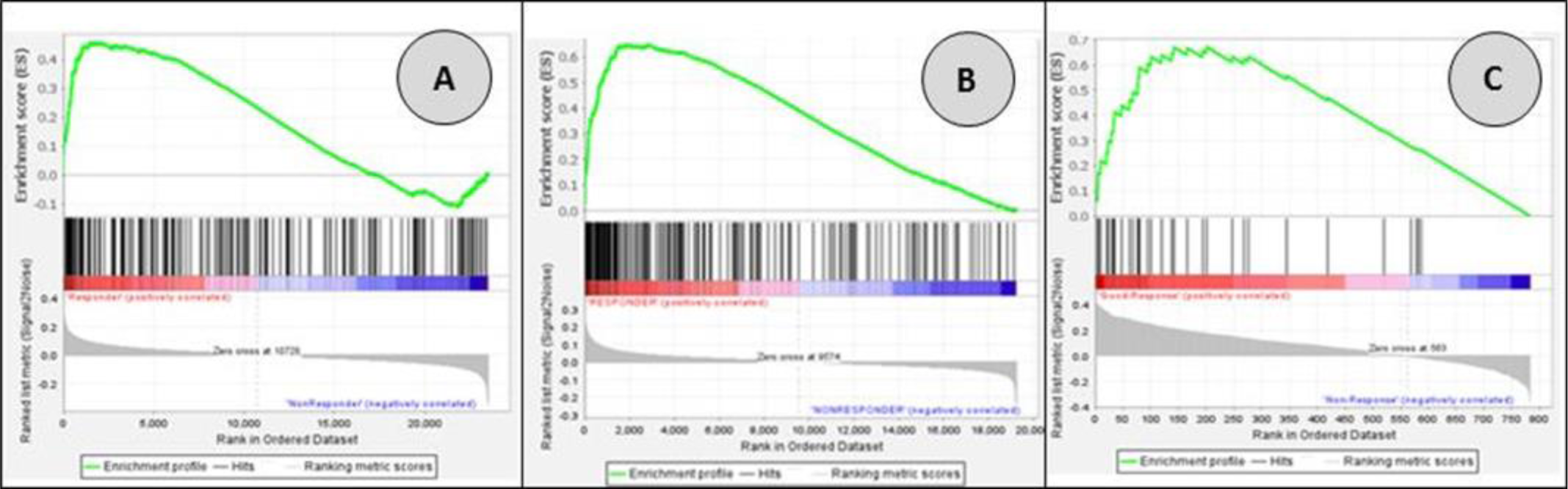
**GSEA enrichment plots for genes included in Hallmark inflammatory response pathway: GSE45404_570 (A); GSE46862 (B); GSE139255 (C).**

**Table 1.**
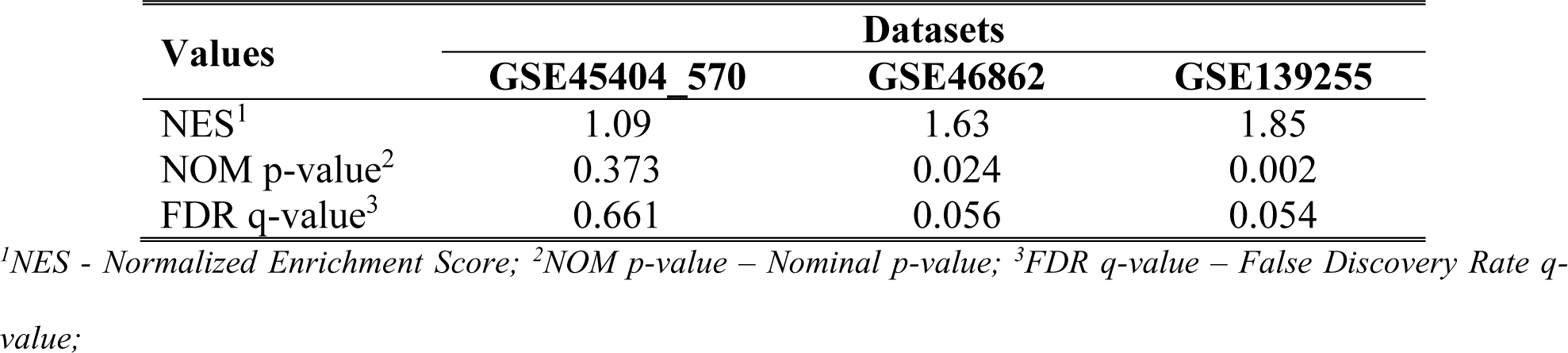
GSEA Hallmark analysis in relation to Hallmark inflammatory response pathway.

The top 100 genes from each database (Supplementary Material 2) were chosen, and overlapped among these three datasets using Venn diagram. The results are presented in Figure 3.

**Figure 3.**
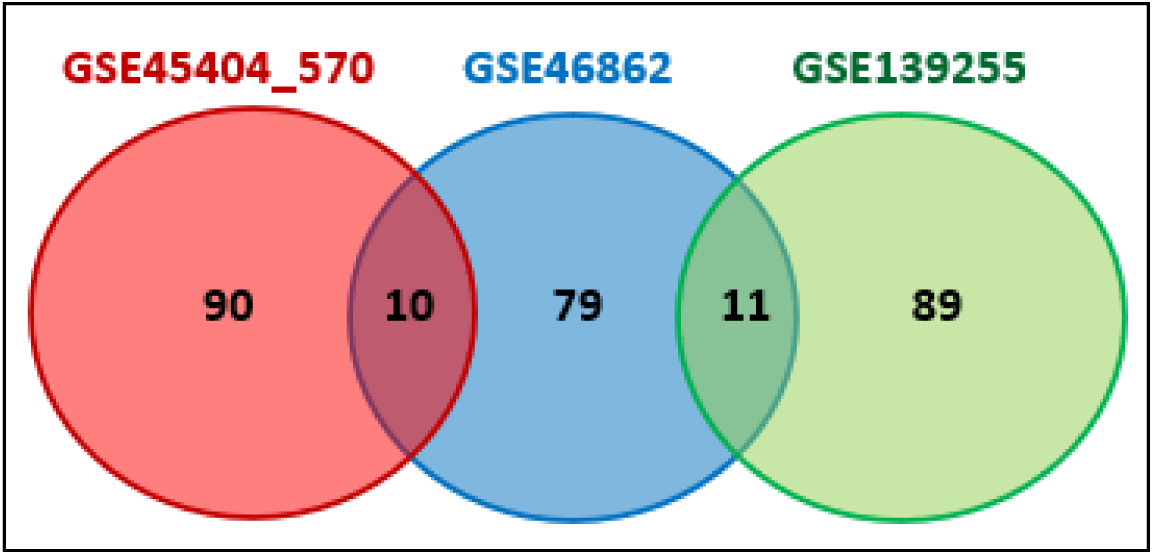
**Venn diagram showing overlapping of inflammation-related genes between three analyzed datasets.**

Our focus was on the genes included in the inflammatory response. As a result of overlapping three datasets, there were 11 genes present in two of them (PLAU, TGFB2, HGF, IL6, CXCL10, CXCL9, IDO1, INHBA, PDGFRB, CYBB, IL24). Statistical significance of these genes among responders and non-responders in all three datasets was examined and the results are presented in Table 2.

**Table 2.** Comparison between responders and non-responders within analyzed datasets in relation to expression of selected genes.

| Gene |  | Data Sets |  |  |  |  |  |  |  |  |
| --- | --- | --- | --- | --- | --- | --- | --- | --- | --- | --- |
|  |  | GSE45404_570 |  |  | GSE46862 |  |  | GSE139255 |  |  |
|  |  | *R* | *NonR* | *Test* * | *R* | *NonR* | *Test* * | *R* | *NonR* | *Test* * |
| *N* (%) |  | 19/42 (45.2%) | 23/42 (54.8%) | - | 28/69 (40.6%) | 41/69 (59.4%) | - | 89/156 (57.1%) | 67/156 (42.9%) | - |
| *PLAU* | Mean (SD) | 7.6 (0.6) | 7.5 (0.9) | ns | 8.1 (0.9) | 7.6 (0.9) | *p*<0.05 | 1967.0 (2152.2) | 766.5 (637.0) | *p*<0.01 |
|  | Median (Range) | 7.7 (6.2-8.5) | 7.4 (6.0-9.5) |  | 8.3 (5.8-9.8) | 7.8 (5.6-9.3) |  | 1351.0 (75.0-9876.0) | 604.7 (88.4-3027.0) |  |
| *TGFB2* | Mean (SD) | 5.0 (0.3) | 5.2 (0.3) | ns | 5.5 (0.7) | 5.1 (0.6) | *p*<0.05 | 64.9 (50.9) | 35.9 (25.9) | *p*<0.01 |
|  | Median (Range) | 5.1 (4.4-5.7) | 5.1 (4.5-5.7) |  | 5.5 (4.3-6.5) | 5.1 (3.7-6.7) |  | 51.6 (1.7-261.9) | 30.8 (3.3-114.8) |  |
| *HGF* | Mean (SD) | 3.8 (0.3) | 3.8 (0.3) | ns | 5.6 (0.9) | 5.2 (0.8) | ns | 332.6 (371.5) | 166.0 (104.4) | *p*<0.01 |
|  | Median (Range) | 3.8 (3.5-4.4) | 3.8 (3.4-4.4) |  | 5.6 (4.0-7.6) | 5.0 (3.5-6.7) |  | 224.6 (22.2-2733) | 160.0 (30.7-540.1) |  |
| *IL6* | Mean (SD) | 6.0 (1.3) | 5.6 (1.1) | ns | 5.7 (1.4) | 4.9 (1.1) | *p*<0.01 | 432.9 (996.6) | 104.3 (200.9) | *p*<0.01 |
|  | Median (Range) | 5.8 (4.0-8.5) | 5.4 (4.2-8.8) |  | 5.7 (4.0-9.7) | 4.5 (3.8-8.8) |  | 90.7 (1.7-5968.0) | 40.7 (1.1-1207.0) |  |
| *CXCL10* | Mean (SD) | 7.9 (1.4) | 6.7 (1.6) | *p*<0.05 | 6.4 (1.3) | 5.8 (1.1) | ns | - | - | NA |
|  | Median (Range) | 7.8 (5.5-10.3) | 6.6 (3.6-10.4) |  | 6.2 (4.1-9.5) | 5.6 (4.0-9.5) |  | - | - |  |
| *CXCL9* | Mean (SD) | 6.8 (1.6) | 5.6 (1.6) | *p*<0.05 | 5.5 (1.0) | 5.0 (0.8) | *p*<0.05 | - | - | NA |
|  | Median (Range) | 6.7 (3.4-9.2) | 5.6 (3.2-9.8) |  | 5.1 (4.3-8.6) | 4.8 (4.1-8.5) |  | - | - |  |
| *IDO1* | Mean (SD) | 6.5 (1.5) | 5.8 (1.2) | ns | 5.9 (1.0) | 5.4 (1.2) | *p*<0.05 | - | - | NA |
|  | Median (Range) | 6.2 (4.5-10.6) | 5.6 (4.2-8.8) |  | 5.7 (3.7-8.2) | 5.2 (3.6-8.5) |  | - | - |  |
| *INHBA* | Mean (SD) | 8.0 (0.8) | 7.9 (1.4) | ns | 6.8 (1.1) | 6.2 (0.9) | ns | 539.0 (768.1) | 240.6 (217.8) | *p*<0.01 |
|  | Median (Range) | 7.9 (6.7-9.4) | 8.1 (5.4-10.7) |  | 6.6 (5.2-9.2) | 6.4 (4.3-8.3) |  | 295.1 (25.2-4582.0) | 172.2 (17.7-1138.0) |  |
| *PDGFRB* | Mean (SD) | 6.9 (0.6) | 6.8 (1.0) | ns | 7.6 (0.8) | 7.1 (0.7) | *p*<0.01 | 1064.0 (894.3) | 629.1 (614.6) | *p*<0.01 |
|  | Median (Range) | 6.7 (6.1-8.2) | 6.9 (4.7-8.9) |  | 7.5 (6.5-9.2) | 7.1 (5.6-8.7) |  | 781.7 (43.7-3629.0) | 470.9 (59.3-3896.0) |  |
| *CYBB* | Mean (SD) | 5.3 (0.6) | 4.7 (0.5) | *p*<0.01 | 8.1 (0.9) | 7.5 (1.1) | *p*<0.01 | - | - | NA |
|  | Median (Range) | 5.0 (4.5-6.4) | 4.6 (3.7-5.5) |  | 8.4 (5.5-9.9) | 7.7 (4.6-9.6) |  | - | - |  |
| *IL24* | Mean (SD) | 5.3 (1.2) | 5.5 (1.4) | ns | 4.8 (0.9) | 4.4 (0.8) | ns | 150.9 (241.4) | 59.7 (66.8) | ns |
|  | Median (Range) | 5.3 (3.3-7.9) | 5.4 (3.4-8.8) |  | 4.6 (3.7-7.1) | 4.1 (3.5-7.1) |  | 56.5 (1.0-1321.0) | 32.7 (2.2-391.9) |  |

*ns* - not statistically significant; *R* - responder; *NonR* - non-responder; \*Wilcoxon rank sum test; NA – not available (without data in dataset); *PLAU* - Plasminogen Activator, *Urokinase*; *TGFB2* - Transforming Growth Factor Beta 2; *HGF* - hepatocyte growth factor; *IL6* - Interleukin-6; *CXCL10* - C-X-C Motif Chemokine Ligand 10; *CXCL9* - C-X-C Motif Chemokine Ligand 9; *IDO1* - Indoleamine 2,3-dioxygenase-1; *INHBA* - Inhibin Subunit Beta A; *PDGFRB* - Platelet derived growth factor receptor beta; *CYBB* - Cytochrome B-245 Beta Chain; *IL24* - Interleukin 24;

None of the selected genes were found to be statistically significant in all three datasets. In order to validate the potential of gene expression to predict treatment outcome, genes included in the Hallmark inflammatory response pathway (IL6, CXCL9, CYBB) were chosen, along with a gene which had promising potential according to literature search (IDO1)^37, 38^. After the connection among selected genes was checked using Cytoscape network, it was found that three of them (IL6, CXCL9, CYBB) were part of a pathway related to avoiding immune detection.

In order to explore the significance of *in silico* obtained results, the expression of candidate genes was analyzed in the cohort of LARC patients from the Institute for Oncology and Radiology of Serbia. Patients, disease, treatment and outcomes characteristics are presented in Table 3. The majority of patients had T3 stadium and N positive disease. One third of patients were female. All patients completed the planned nCRT. Operative treatment was conducted in 63 patients, and the pathohistological complete response rate was 20.6 % (Table 3). Twelve patients with distally located tumor and complete clinical response were involved in a “watch and wait” approach. One patient had to be excluded from the hematological ratios analysis, because of his chronic lymphocytic leukemia and its influence to the parameters of this analysis.

**Table 3.**
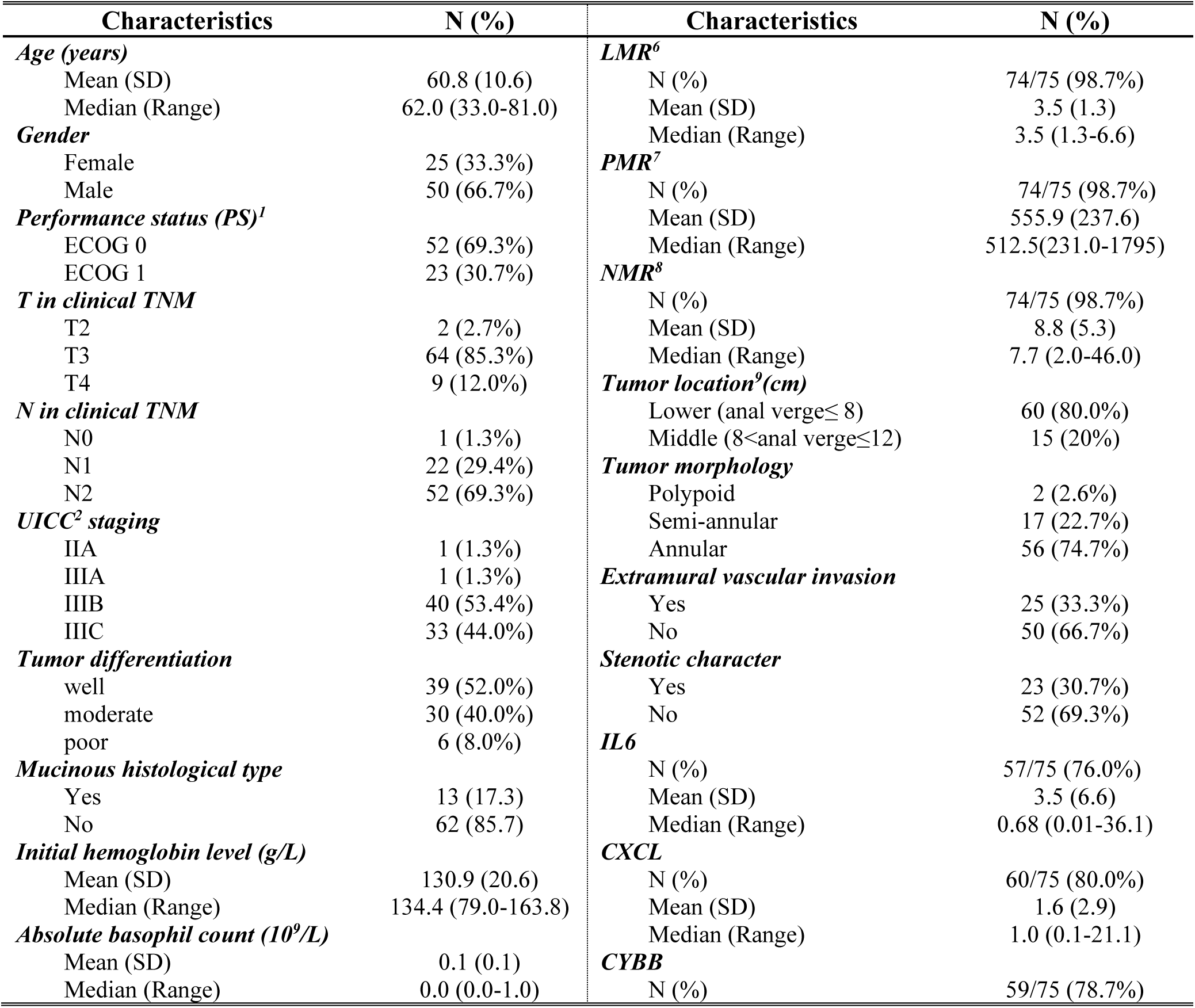
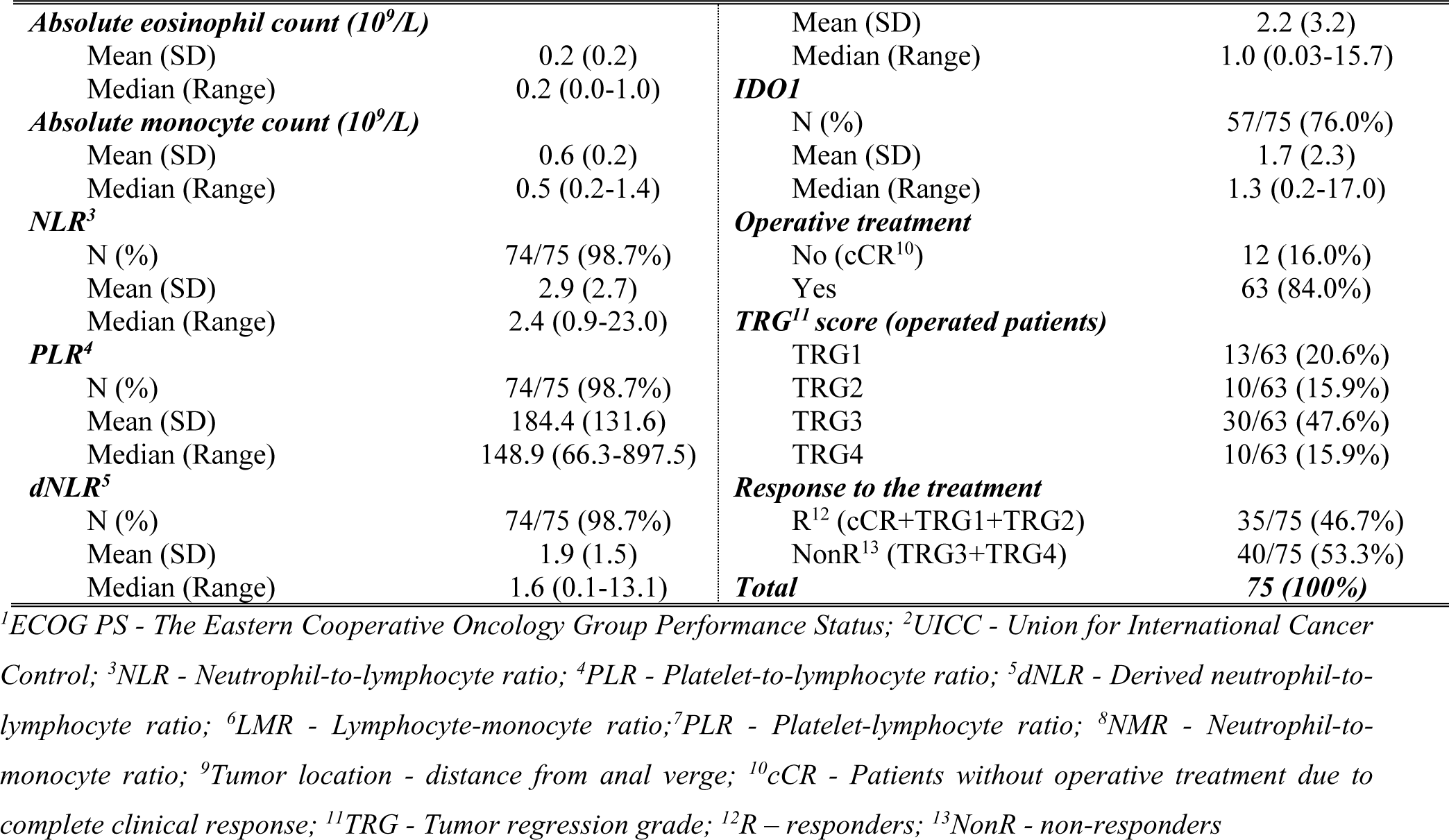
Patients’, disease, treatment and outcomes characteristics.

Correlation of clinical evaluation and pathological examination as a gold standard in a group of patients where operative treatment was conducted is presented in Table 4. Using disease prevalence of 79.4% the sensitivity, specificity, positive predictive value, negative predictive value and predictive accuracy were calculated (Table 5).

**Table 4.** Correlation of clinical and pathological CR within a group of patients where operative treatment was conducted.

|  |  | Pathological assessment of disease presence |  |  |
| --- | --- | --- | --- | --- |
|  |  | Yes | No | Total |
| Clinical assessment of disease presence | Yes | 46 (73.02%) | 3 (4.76%) | 49 (77.78%) |
|  | No | 4 (6.35%) | 10 (15.87%) | 14 (22.22%) |
|  | Total | 50 (79.37%) | 13 (20.63%) | 63 (100%) |

**Table 5.** Sensitivity, specificity, positive predictive value, negative predictive value and predictive accuracy of clinical evaluation for prediction of disease status using pathological examination as a gold standard.

| Characteristics | Clinical evaluation |
| --- | --- |
| *Sensitivity (95% CI)* | 92.0% (80.8-97.8%) |
| *Specificity (95% CI)* | 76.9% (46.2-95.0%) |
| *PPV (95% CI)* | 93.9% (85.0-97.6%) |
| *NPV (95% CI)* | 71.4% (48.2-87.0%) |
| *Predictive accuracy (95% CI)* | 88.9% (78.4-95.4%) |

*CI – confidence interval; NPV – negative predictive value; PPV -positive predictive value*

Research interest was the comparisons between responders (comprised 35 patients) and non-responders (included 40 patients) (Table 6). Initial T and N stadium of disease were not significantly different between these two groups. Patients with poorly differentiated tumors and those with mucinous histological type responded to treatment significantly worse than patients with well or moderate tumor differentiation and those without mucinous type (p<0.05 and p<0.01, respectively). According to MRI findings, non-responders presented more often with extramural vascular invasion (EMVI) (p<0.05). Among hematological parameters, significance was found for absolute basophil, eosinophil and monocyte counts, dNLR and NMR.

**Table 6.** Comparison of characteristics of responders and non-responders to neoadjuvant chemoradiotherapy.

| Characteristic | The response to treatment |  |  |
| --- | --- | --- | --- |
|  | Responders | Non-responders | Wilcoxon rank sum test |
| **Age (years)** |  |  |  |
| Mean (SD) | 61.5 (10.7) | 60.3 (10.6) | *ns* |
| Median (Range) | 63.0 (38.0-81.0) | 62.0 (33.0-76.0) |  |
| **Gender** |  |  |  |
| Male | 25 (71.4%) | 25 (62.5%) | *ns\** |
| Female | 10 (28.6%) | 15 (37.5%) |  |
| **T in clinical TNM** |  |  |  |
| T2 | 2 (5.7%) | 0 (0%) | *ns<sup>#</sup>* |
| T3 | 29 (82.9%) | 35 (87.5%) |  |
| T4 | 4 (11.4%) | 5 (12.5%) |  |
| **N in clinical TNM** |  |  |  |
| N0 | 1 (2.9%) | 0 (0%) | *ns<sup>#</sup>* |
| N1 | 13 (37.1%) | 9 (22.5%) |  |
| N2 | 21 (60.0%) | 31 (77.5%) |  |
| **UICC staging** |  |  |  |
| IIA+ IIIA+ IIIB | 22 (62.9%) | 20 (50.0%) | *ns\** |
| IIIC | 13 (37.1%) | 20 (50.0%) |  |
| **Tumor differentiation** |  |  |  |
| Well and moderate | 35 (100%) | 34 (85%) | $p<0.05^{\#}$ |
| Poor | 0 (0%) | 6 (15%) |  |
| **Mucinous histological type** |  |  |  |
| No | 35 (100%) | 27 (67.5%) | $p<0.01^{\#}$ |
| Yes | 0 (0%) | 13 (32.5%) |  |
| **Extramural vascular invasion** |  |  |  |
| No | 28 (80%) | 22 (55%) | $p<0.05^*$ |
| Yes | 7 (20%) | 18 (45%) |  |
| **Tumor morphology** |  |  |  |
| Polypoid and semi-annular | 14 (40.0%) | 5 (12.5%) | $p<0.01^*$ |
| Annular | 21 (60.0%) | 35 (87.5%) |  |
| **Stenotic character** |  |  |  |
| No | 29 (82.9%) | 23 (57.5%) | $p<0.05^*$ |
| Yes | 6 (17.1%) | 17 (42.5%) |  |
| **Absolute basophil count** |  |  |  |
| Mean (SD) | 0.03 (0.04) | 0.08 (0.16) | $p<0.01$ |
| Median (Range) | 0.02 (0-0.1) | 0.1 (0-1.0) |  |
| **Absolute eosinophil count** |  |  |  |
| Mean (SD) | 0.17 (0.12) | 0.27 (0.22) | $p<0.05$ |
| Median (Range) | 0.1 (0-0.53) | 0.2 (0-1.0) |  |
| **Absolute monocyte count** |  |  |  |
| Mean (SD) | 0.52 (0.18) | 0.63 (0.21) | $p<0.01$ |
| Median (Range) | 0.5 (0.2-1.1) | 0.6 (0.4-1.4) |  |
| **Neutrophil-to-lymphocyte ratio** |  |  |  |
| Mean (SD) | 3.38 (3.62) | 2.56 (1.5) | $ns$ |
| Median (Range) | 2.5 (1.17-23.0) | 2.27 (0.93-7.46) |  |
| **Platelet-to-lymphocyte ratio** |  |  |  |
| Mean (SD) | 184.8 (141.5) | 184.0 (123.9) | $ns$ |
| Median (Range) | 144.4 (72.3-897.5) | 150.0 (66.3-681.1) |  |
| **dNLR<sup>§</sup>** |  |  |  |
| Mean (SD) | 2.21 (2.02) | 1.61 (0.73) | $p<0.05$ |
| Median (Range) | 1.81 (0.84-13.14) | 1.49 (0.09-3.37) |  |
| **Neutrophil-to-monocyte ratio** |  |  |  |
| Mean (SD) | 10.3 (7.01) | 7.44 (2.48) | $p<0.01$ |
| Median (Range) | 9.18 (4.56-46.0) | 7.0 (2.0-15.5) |  |
| **IL6** |  |  |  |
| N (%) | 28/35 (80.0%) | 29/40 (72.5%) | $ns$ |
| Mean (SD) | 4.3 (8.6) | 2.8 (4.0) |  |
| Median (Range) | 0.4 (0.01-36.1) | 1.4 (0.05-16.8) |  |
| **CYBB** |  |  |  |
| N (%) | 29/35 (82.8%) | 30/40 (75.0%) | $ns$ |
| Mean (SD) | 2.1 (2.8) | 2.2 (3.6) |  |
| Median (Range) | 1.0 (0.03-10.3) | 1.1 (0.05-15.7) |  |
| **CXCL9** |  |  |  |
| N (%) | 29/35 (82.8%) | 31/40 (77.5%) | $ns$ |
| Mean (SD) | 1.1 (0.7) | 2.1 (3.9) |  |
| Median (Range) | 0.9 (0.3-2.4) | 1.2 (0.1-21.1) |  |
| **IDO1** |  |  |  |
| N (%) | 28/35 (80.0%) | 29/40 (72.5%) | $ns$ |
| Mean (SD) | 1.5 (1.1) | 1.9 (3.1) |  |
| Median (Range) | 1.4 (0.2-3.6) | 1.3 (0.2-17.0) |  |
| **Total** | **35 (100%)** | **40 (100%)** | **-** |

\*Pearson $\chi^2$ Test; <sup>#</sup>Fisher Exact Test; $ns$ - not statistically significant; <sup>§</sup>dNLR-Derived neutrophil-to-lymphocyte ratio

In the whole patient group, there was no significant correlation between *in silico* selected genes (IL6, CYBB, CXCL9, IDO1) and response to treatment. On the other hand, when comparison between patients where pCR (TRG1) was detected and those who responded the worst (TRG4), statistical significance was found based on IDO1 expression (Wilcoxon rank sum test, *p*=0.036) (Supplementary Table 1).

Next, ROC analysis was performed and it revealed the optimal cut-off values for absolute basophil, eosinophil and monocyte counts and NMR, above/below which the possibility of achieving favorable response after nCRT increased significantly (Table 7, Figure 4). The optimal cut-off value, which might distinguish patients with and without good response was not found only for dNLR.

**Figure 4.**
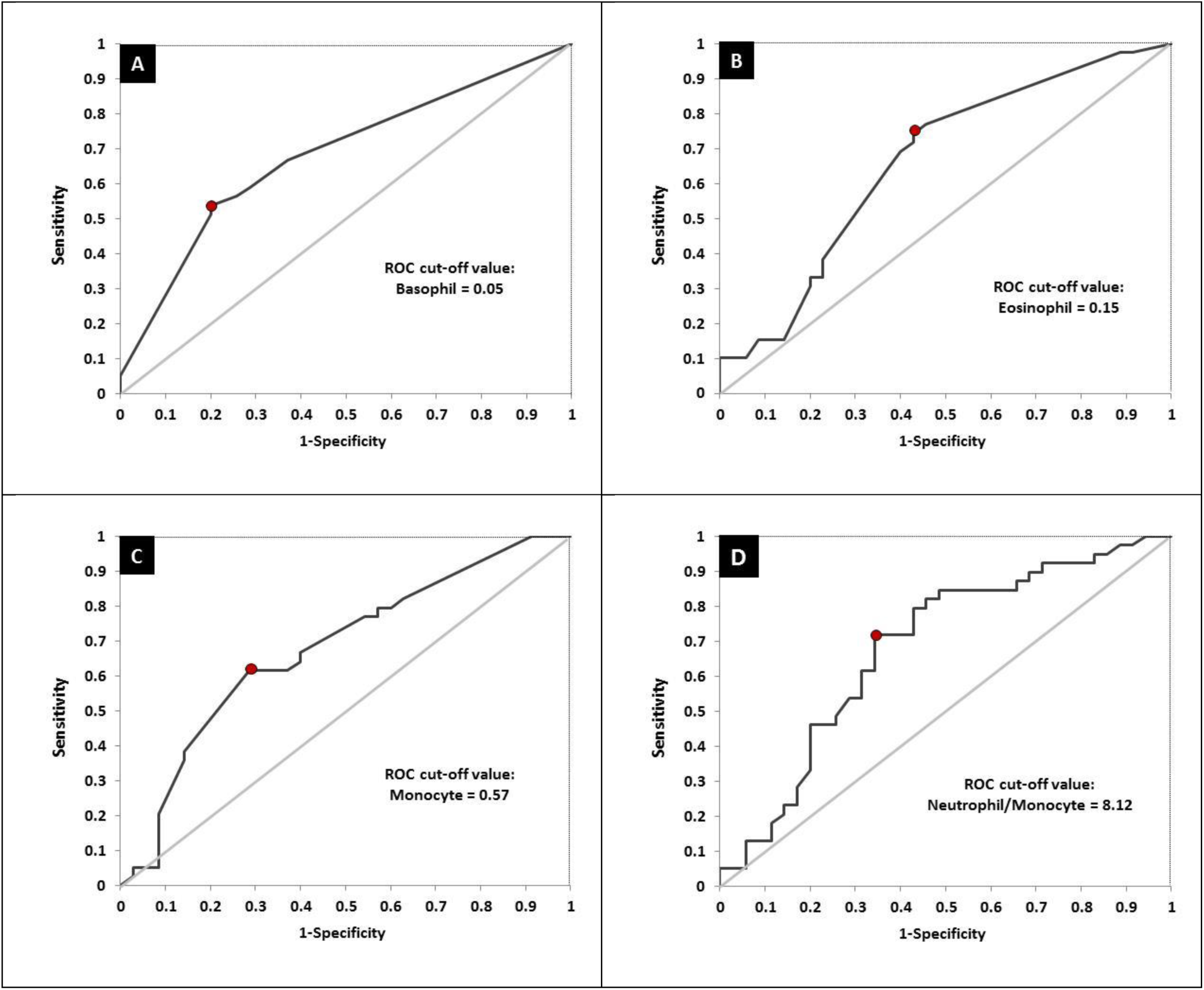
**ROC curves for the absolute basophil count (A), absolute eosinophil count (B), absolute monocyte count (C) and NMR (D) in relation to response to treatment.**

**Table 7.** Results of the ROC analysis for NMR, dNLR, absolute basophil, eosinophil and monocyte counts, and relevant events.

| Characteristics | Absolute count |  |  | dNLR | NMR |
| --- | --- | --- | --- | --- | --- |
|  | Basophil | Eosinophil | Monocyte |  |  |
| **AUC ROC<sup>a</sup>** | 68.2% | 66.1% | 67.8% | 62.9% | 68.2% |
| **(95% CI)** | (56.5-79.9%) | (53.7-78.6%) | (55.5-80.1%) | (50.0-75.8%) | (55.6-80.8%) |
| **Likelihood ratio test<sup>b</sup>** | *p*<0.01 | *p*<0.05 | *p*<0.05 | *ns* | *p*<0.01 |
| **ROC-cut-off value<sup>c</sup>** | 0.05 | 0.15 | 0.57 | - | 8.12 |
| **Sensitivity** | 53.8% | 74.4% | 61.5% | - | 71.8% |
| **(95% CI)** | (38.5-69.2%) | (59.0-87.2%) | (46.2-76.9%) | - | (56.4-84.6%) |
| **Specificity** | 80.0% | 57.1% | 71.4% | - | 65.7% |
| **(95% CI)** | (65.7-91.4%) | (40.0-71.4%) | (57.1-85.7%) | - | (48.6-80.0%) |

<sup>a</sup>Area Under the ROC curve (DeLong's method); <sup>b</sup>Likelihood ratio test for AUC ROC; <sup>c</sup>Value with maximum sensitivity and specificity; *ns* - not statistically significant

Afterwards, differences between responders and non-responders according to the cut-off values obtained by ROC analysis were examined (Table 8). According to the achieved cut-off values a statistically significant difference in the response was confirmed for the initial basophil, eosinophil and monocyte counts (p < 0.01 for all variables). Initial higher level of these parameters (greater than 0.05, 0.15, 0.57 respectively) were associated with unfavourable responses.

**Table 8.** The value of NMR, dNLR, absolute basophil, eosinophil and monocyte counts in prediction the response to nCRT.

| Parameters<br>(ROC cut-off value) | Response to nCRT |  |  |
| --- | --- | --- | --- |
| | responders | non-responders | Pearson $\chi^2$ test |
| ***Absolute basophil count*** |  |  |  |
| ≤ 0.05 | 28 (80.0%) | 18 (45.0%) | $p < 0.01$ |
| > 0.05 | 7 (20.0%) | 22 (55.0%) |  |
| ***Absolute eosinophil count*** |  |  |  |
| ≤ 0.15 | 20 (57.1%) | 10 (25.0%) | $p < 0.01$ |
| > 0.15 | 15 (42.9%) | 30 (75.0%) |  |
| ***Absolute monocyte count*** |  |  |  |
| ≤ 0.57 | 25 (71.4%) | 15 (37.5%) | $p < 0.01$ |
| > 0.57 | 10 (28.6%) | 25 (62.5%) |  |
| ***Neutrophil-to-monocyte ratio*** |  |  |  |
| ≤ 8.12 | 12 (34.3%) | 28 (70.0%) | $p < 0.05$ |
| > 8.12 | 32 (65.7%) | 12 (30.0%) |  |
| **Total** | **35 (46.7%)** | **40 (53.3%)** | **-** |

Significant variables from the analyses were then used for the construction of a logistic regression model. The UICC staging was included as parameter which unit T and N stadium of disease and has high clinical importance. Finally, the model comprised ten variables: UICC staging, tumor differentiation, mucinous histological type, tumor morphology, stenotic character, extramural vascular invasion, as well as NMR, absolute basophil, eosinophil and monocyte counts (Table 9). After univariate anayses were conducted, the extremely high OR values were observed for tumor differentiation and mucinous histological type categories. These values were in correlation with the fact that all patients with mucinous histological type and/or poorly differentiated tumor had achieved bad response. Previously mentioned parameters as well as UICC staging were excluded after univariate analyses. The final model included tumor morphology, NMR, absolute basophil, eosinophil, and monocyte counts.

**Table 9.** Logistic regression analysis of the response to nCRT.

| Characteristic | Logistic regression |  |  |  |
| --- | --- | --- | --- | --- |
|  | Univariate |  | Multivariate |  |
|  | Odds Ratio (95%CI) | Wald test | Odds Ratio (95%CI) | Likelihood Ratio test |
| ***UICC staging<sup>s</sup>*** |  |  |  |  |
| IIIC vs. IIA+ IIIA+ IIIB | 1.69 (0.67-4.26) | $p=0.265$ | - | - |
| ***Tumor differentiation*** |  |  |  |  |
| Poor vs. Well and moderate | 43.8*10<sup>6</sup> (0-∞) | $p=0.991$ | - | - |
| ***Mucinous histological type*** |  |  |  |  |
| Yes vs. No | 14.99*10<sup>7</sup> (0-∞) | $p=0.992$ | - | - |
| ***Tumor morphology*** |  |  |  |  |
| Annular vs. Polypoid and semi-annular | 4.67 (1.47-14.82) | $p=0.009$ | 10.11 (1.81-56.39) | $p^{\#}=0.008$ |
| ***Stenotic character*** |  |  |  |  |
| Yes vs. No | 3.57 (1.21-10.52) | $p=0.021$ | - | $p=0.230$ |
| ***Extramural vascular invasion*** |  |  |  |  |
| Yes vs. No | 3.27 (1.16- 9.23) | $p=0.025$ | - | $p=0.131$ |
| ***Absolute basophil count*** |  |  |  |  |
| > 0.05 vs. ≤ 0.05 | 4.89 (1.73-13.78) | $p=0.003$ | 4.55 (1.21-17.13) | $p^{\#}=0.025$ |
| ***Absolute eosinophil count*** |  |  |  |  |
| > 0.15 vs. ≤ 0.15 | 4.00 (1.50-10.66) | $p=0.005$ | 3.86 (1.09-13.71) | $p^{\#}=0.037$ |
| ***Absolute monocyte count*** |  |  |  |  |
| > 0.57 vs. ≤ 0.57 | 4.17 (1.57-11.03) | $p=0.004$ | 3.46 (1.01-11.89) | $p^{\#}=0.049$ |
| ***NMR<sup>ε</sup>*** |  |  |  |  |
| ≤ 8.12 vs. > 8.12 | 4.47 (1.69-11.82) | $p=0.003$ | 6.38 (1.74-23.39) | $p^{\#}=0.005$ |

<sup>s</sup>UICC - Union for International Cancer Control; <sup>ε</sup>NMR - Neutrophil-to-monocyte ratio; <sup>#</sup>Wild test.

The numerical variables that remained significant in the multivariate analysis were utilized to create eleven different composite scores. These scores were calculated using various combinations of the significant variables (Supplementary Table 2). The best predictive power was observed when the initial eosinophil, basophil, and monocyte counts were combined (Figure 5). The changes in the false negative and true positive rates for the top three composite scores with respect to different cut-off values of these three scores are shown in Figure 6.

**Figure 5.**
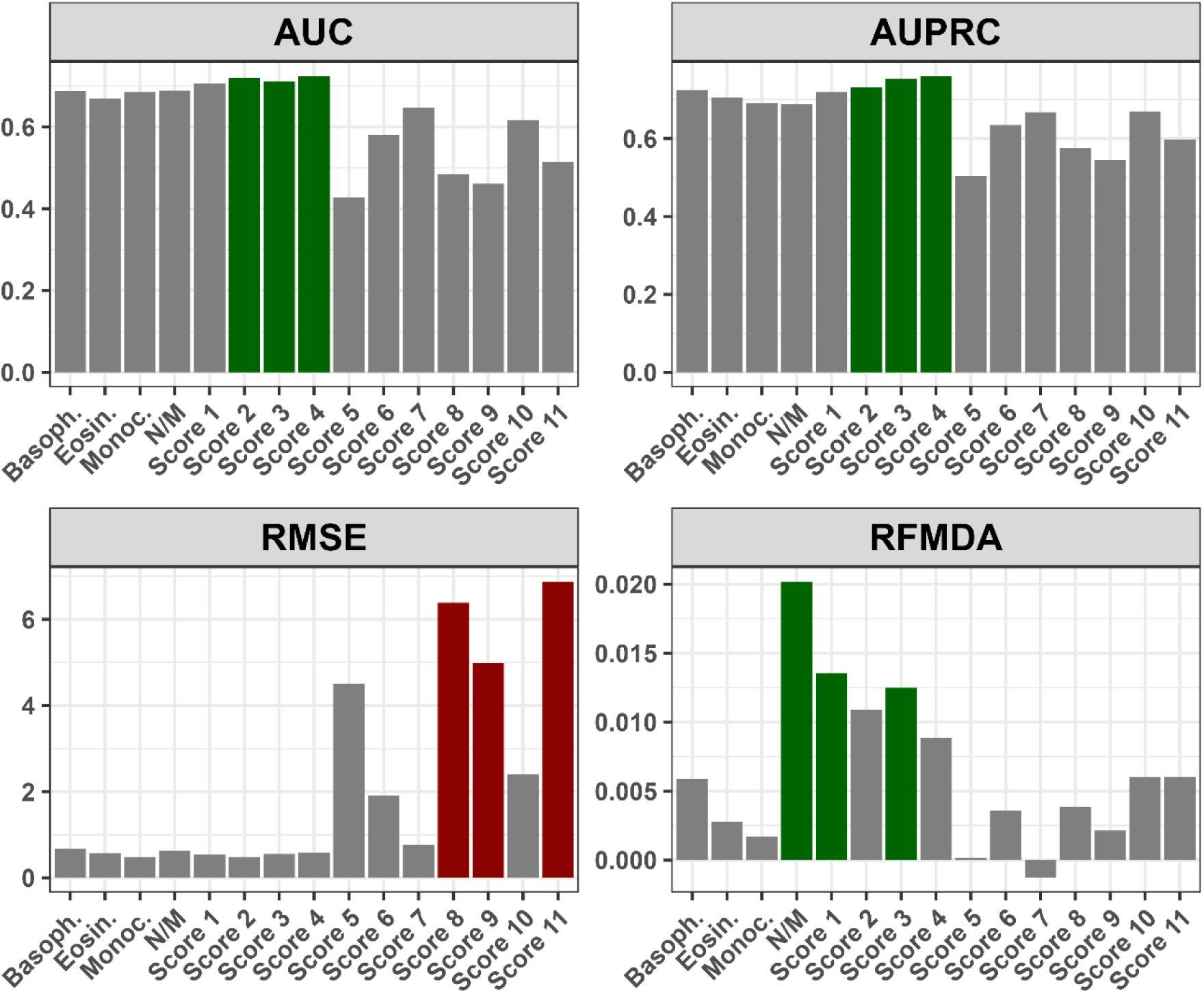
**Performance of the composite scores with respect to various metrics: AUC - Area Under Curve; AUPRC - Area Under Precision-Recall Curve; RMSE - Root Mean Square Error; RFMDA - Random Forest Mean Decrease in Accuracy; Basoph. - Absolute basophil count; Eosin. - Absolute eosinophil count; Monoc. - Absolute monocyte count; N/M - Neutrophil-to-monocyte ratio; Score 1 - Absolute basophil + eosinophil count; Score 2 - Absolute basophil + monocyte count; Score 3 - Absolute eosinophil + monocyte count; Score 4 - Absolute basophil + eosinophil + monocyte count; Score 5 - Neutrophil-to-monocyte ratio + Absolute monocyte count; Score 6 - Neutrophil-to-monocyte ratio + Absolute eosinophil count; Score 7 - Neutrophil-to-monocyte ratio + Absolute basophil count; Score 8 - Neutrophil-to-monocyte ratio + Absolute monocyte + eosinophil count; Score 9 - Neutrophil-to-monocyte ratio + Absolute monocyte + Absolute basophil count; Score 10 - Neutrophil-to-monocyte ratio + Absolute eosinophil + basophil count; Score 11 - Neutrophil-to-monocyte ratio + Absolute monocyte + eosinophil count + basophil count;**

**Figure 6.**
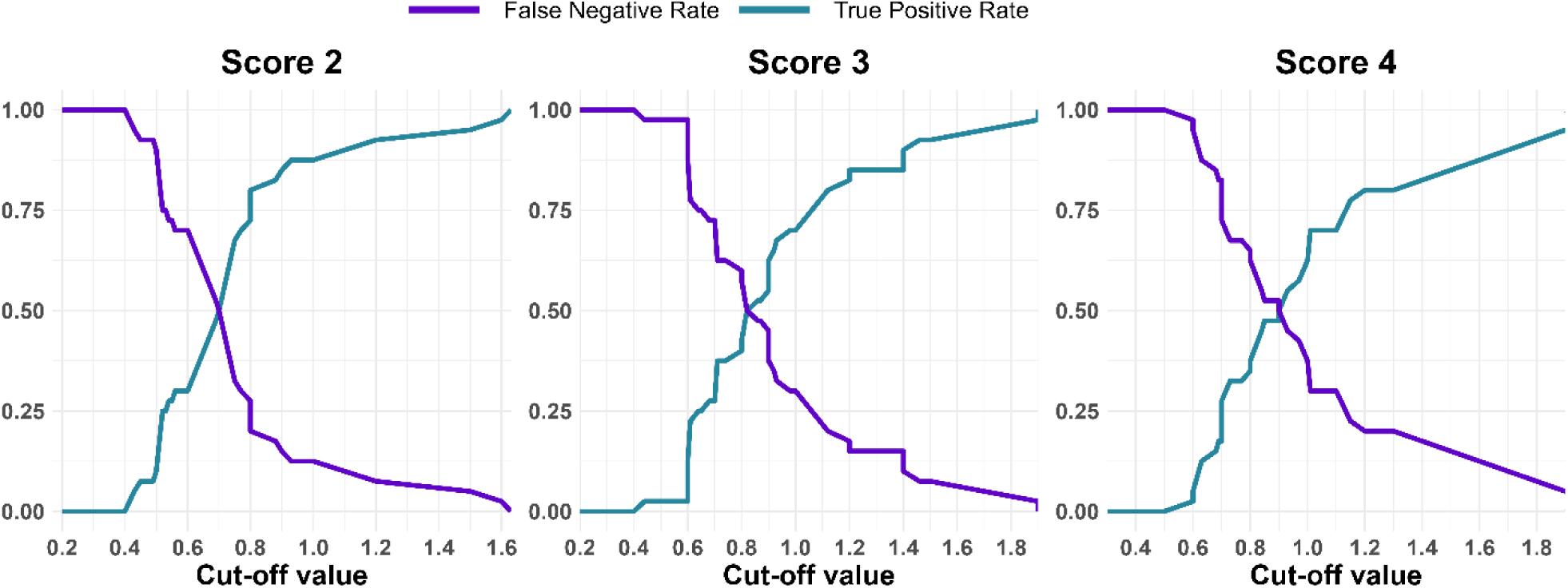
**Relationship between False Negative and True Positive Rates for Top Three Composite Scores at Different Cut-off Values: Score 2 - Absolute basophil + monocyte count; Score 3 - Absolute eosinophil + monocyte count; Score 4 - Absolute basophil + eosinophil + monocyte count;**

## Discussion

The optimal time for assessment of tumor response after nCRT, time for surgery, and how to profile the best candidates for the „watch and wait” approach is still unknown. In this study, we aimed to select patients who would benefit the most from an increase of RT dose and waiting periods longer than 6 weeks after nCRT completion according to initial clinical, pathological, radiological, and hematological parameters, as well as inflammation-related genetic biomarkers chosen by *in silico* analysis. The identification of these predictive clinical and molecular markers would enable the intensification of treatment in selected groups of patients. Better selection of patients with a higher probability of a favorable response to neoadjuvant treatment would contribute to the reduction of morbidity, while improving survival and local control of the disease. On the other hand, patients where a good response to neoadjuvant treatment is not expected would be candidates for other treatment modalities in the initial approach, such as induction polychemotherapy, application of target therapy (e.g. epidermal growth factor receptor inhibitors) or surgery without delay after completion of neoadjuvant treatment.

In some cases, pelvic MRI scan performed at 8th week after nCRT completion cannot clearly distinguish residual tumor due to post treatment changes and still probably did not achieve the maximum response. The sensitivity of clinical evaluation, according to our results, was 92%. It refers to high probability that disease evaluation will indicate an incomplete response when viable tumor cells are present, which is confirmed with pathohystological examination as a gold standard. Therefore, the combination of MRI scan and proctoscopy examination is beneficial when it comes to a group of patients who still have residual disease after nCRT, at which point operative treatment is indicated. On the other hand, lower specificity and negative predictive value (NPV) (76.92%, 71.39%, respectively) suggest that this kind of evaluation is not selective enough for patients who are candidates for the “watch and wait” approach. This method is particularly important in the case of distally located rectal cancer when abdominoperineal resection is the only option. In our study, the majority of patients had distant located tumor (80%). The only way to confirm CR after nCRT is strict follow-up with reevaluation every 2-3 months in the first 2 years after treatment completion, followed by continuation of the protocols ^39^. Evidence suggest that in the case of local regrowth, salvage surgery can be done in 95% of patients, which indicate the safety of this approach ^10^. However, when near CR is found at the first assessment, the protocols are not well established yet. It is well known that prolongation of period after nCRT completion is associated with higher pCR rate ^40^. In the case when primary response 6-8 weeks after treatment completion is close to cCR, it is beneficial for patients who are not candidates for sphincter preservation surgery to delay surgery with one more clinical assessment after 8-12 weeks in order to achieve the maximum response. Simpson at al. reported local regrowth on repeated assessment for 37% of patients whose response was defined as near CR ^41^. Another article which investigated the role of prolongation of period after nCRT in order to achieve the maximum response, found that 90% of patients with initial near CR at the first assessment were found to be cCR at the reassessment after 6-12 weeks ^42^. On the other hand, delaying surgery in order to achieve better response is associated with a higher probability of distant metastases ^43^. This fact can be related to local regrowth, but it has not been proved yet.

These circumstances stress the necessity of additional parameters which can guide the selection of patients who can be expected to achieve a complete response. Molecular markers in combination with good MRI and rigid proctoscopy examination may allow longer delays in surgery and one more pelvic MRI scan after 8-12 weeks. In this study, we aimed to investigate some genetic factors that were found to be promising candidates using *in silico* methods of previously published datasets. However, statistical significance between responders and non-responders in relation to expression of selected genes (*IL6, CYBB, CXCL9, IDO1*) was not reached. When comparisons were made in the subgroup of patients who were operated, a significantly higher expression of *IDO1* (*p*<0.05) was found for TRG1 compared to TRG4. IDO1 is critical for tryptophan metabolism, and is regarded to have a significant effect on the modulation of T-cell behavior and differentiation of regulatory T-cells. In a previous study which explored IDO1 expression using immunohistochemistry in postoperative specimens, the relation to pathological response was not found (*p*=0.44). The same study showed that higher expression of IDO1 was associated with worse prognosis ^38^. However, another study exploring nodal-positive LARC revealed that high IDO1 expression in specimens after nCRT completion was associated with improved overall survival (OS) ^44^. In our study, all but one patient were nodal-positive and our analyses were conducted on the initial specimens, which enabled us to analyze potential predictive biomarkers.

Concerning liquid biopsy parameters, periodic measurement of markers during patient follow up may also be crucial to prove the absence of the disease as well as for early detection of disease progression. This kind of approach has been investigated in metastatic colorectal cancer and it was shown that periodic sampling of liquid biopsy accompanied with ctDNA levels measurements can be valid for monitoring status of the disease and profile the response to treatment ^45^.

The importance of EMVI as a prognostic factor in LARC setting is well established. By comparison of disease-free survival (DFS) between II and III stadium of disease, it was shown that independent from disease stadium, the presence of EMVI results in the worse prognosis ^46^. The predictive role of EMVI has not been defined yet. Sun et al. found that EMVI status was the only factor by multivariate analysis which influences the response to treatment. The focus of this research was the role of initial MRI characteristics on treatment outcome of T3 LARC patients. Patients with ypT0-2N0 postoperative category were previously defined as good responders ^47^. In our cohort, 33.3% of patients were EMVI positive, and it was shown that they were more likely to have poor response (*p*<0.05). Worse response in EMVI positive group of patients can be connected with tumour hypoxia and consequent radioresistancy, due to the fact that primary mechanism of radiotherapy effectiveness is formation of reactive oxygen radicals. Hypoxia in solid tumors is a well known problem because of insufficient vascularisation of rapid tumor growth. In order to resolve this in our study, we tried to increase the administrated dose per fraction on the gross disease region (2.16 Gy/fraction). By doing this, we attempt to cause cell death related to direct DNA damage caused by radiation, and to overcome lower level of oxygen in some parts of the tumor. By combining pCR rate in group of patients where operative treatment was conducted and patients who were enrolled in “watch and wait“ program, we achieved 33.3% complete response rate. On the other hand, in EMVI positive group of patients, the complete response was achieved for only 16% of them. The option for this group of patients might be further dose escalation using adaptive MRI-guided radiotherapy which had shown potential for higher cCR rates and wider implementation of organ preservation approaches ^48^.

The role of initial basophile count has been previously investigated as a prognostic factor in colorectal cancer. The association between lower basophile level and worse survival as well as aggressive tumor potential has been shown ^49^. To the best of our knowledge, this is the first study to find the predictive role of basophile counts in the rectal cancer settings. Patients with an initial basophile count lower than 0.05 are more likely to achieve good response (*p*<0.01). Similar results were found in advanced gastric cancer, where worse response to programmed death 1 inhibitor (anti-PD-1 inhibitor) plus chemotherapy was in correlation with a higher level of peripheral basophils ^50^.

Comparing literature data on the predictive role of initial eosinophil counts, it has been proposed as a potential predictive marker for immunotherapy in lung cancer, with a higher levels detected in patients with better treatment outcome ^51^. It was also found that higher initial level of eosinophil is connected to more effective outcomes when immunotherapy is administrated together with chemotherapy in advanced melanoma ^52^. In our study, a higher initial eosinophil level is associated with worse response, which might be explained by different treatment modalities and the addition of the radiotherapy component.

Analyzing initial monocyte counts, a predictive role was previously reported in the CRC settings, with higher levels detected in patients with poor outcome ^53^. The same was found in our research where the absolute monocyte levels a higher than 0.57 were related to worse response. The NMR has been investigated in low-risk differentiated thyroid carcinoma as a prognostic factor, and it was found that lower initial level is related to a worse prognosis, which is in relation to our findings ^54^. Our group has previously shown that hematological parameters easily derived and routinely determined by low-cost and minimally invasive methods might be useful in predicting the response to chemoradiotherapy in patients with anal cancer ^7^. Also, we successfully evaluated the role of hematological parameters in predicting the survival and toxicity to specific treatment in the lung cancer setting ^55^.

According to our results, mucinous tumor differentiation was significantly assocciated with poor response (*p*<0.01). The study conducted by Simha et al. also found that presence of mucin is associated with larger residual disease and worse prognosis ^56^. Previously it has also been described that mucinous rectal carcinoma is associated with a unique genetic pattern, including more frequent presence of microsatellite instability (MSI), which is caused by a defect in DNA mismatch repair ^57^. The connection of MSI in rectal carcinoma and poorer prognosis has also been reported ^58^. Bearing it in mind, recently presented preliminary results with focus on usefulness of introduction of the anti-PD-1 inhibitor dostarlimab in patients with mismatch repair–deficient (dMMR) LARC patients can be promising to individualise treatment in this group of patients ^59^.

This study has some limitations. The sample size is relatively low, but has met the criteria of a minimum number of LARC samples taking into account its incidence and population size in Serbia (95% confidence level) ^60^. The evaluation of potential prognostic parameters has not been included, as the enrolled patients are currently under follow-up for long-term outcomes.The predictive model constructed in our study is currently being validated in an independent prospective cohort of patients with LARC treated with nCRT.

## Conclusions

Based on the logistic regression model, important factors associated with favorable response to nCRT were tumor morphology and hematological parameters which can be easily and routinely derived from initial laboratory results (NMR, eosinophile, basophil and monocyte counts) in a minimally invasive manner. Here, we present evidence that a combined score derived by summing the initial absolute counts of basophils, eosinophils, and monocytes holds the highest predictive value and potential clinical utility. Further studies involving larger cohorts are necessary to validate these initial observations.

## Data Availability

The data that support the findings of this study are available upon reasonable request from the corresponding author. The data are not publicly available due to ethics restrictions as their containing information could compromise the privacy of patients.

## Acknowledgements

This study was funded by the Horizon Europe Project STEPUPIORS (Agreement No. 101079217) and the Ministry of Education and Science of the Republic of Serbia (Agreement No. 451-03-47/2023-01/200043).

## Study approval

This study has been approved by the Ethics Committee of the Institute for Oncology and Radiology of Serbia and the Ethics Committee of the Faculty of Medicine, University of Belgrade, Serbia. All patients signed an informed consent. All experiments have been performed in accordance with the Helsinki Declaration of 1975, as revised in 2013.

## Table and Figure legends

**Supplementary Material 1. The results of the differential expression analysis within GSE 139255.**

**Supplementary Material 2. The top 100 genes from each database.**

**Supplementary Table 1. Comparison of genes expression of patients with pCR and those with TRG4 postoperative category.**

**Supplementary Table 2. Performance of the composite scores with respect to various metrics.**

## List of abbreviations

CRC: colorectal cancer
LARC: locally advanced rectal cancer
MRI: magnetic resonance imaging
nCRT: neoadjuvant chemoradiotherapy
pCR: pathologic complete response
cCR: clinical complete response
NCBI GEO: National center for biotechnology information gene expression omnibus
TRG: tumor regression grading
GSEA: gene set enrichment analysis
KEGG: Kyoto encyclopedia of genes and genomes
CT: computed tomography
RT: radiotherapy
VMAT-SIB: volumetric modulated arc therapy-simultaneous integrated boost technique
PR: partial response
FFPE: formaline-fixed paraffin-embedded
EDTA: ethylenediaminetetraacetic acid
CBC: complete blood count
NLR: neutrophil-to-lymphocyte ratio
PLR: platelet-to-lymphocyte ratio
dNLR: derived neutrophil-to-lymphocyte ratio
LMR: lymphocyte-to-monocyte ratio
PMR: platelet-to-monocyte ratio
NMR: neutrophil-to-monocyte ratio
UICC: Union for International Cancer Control
ECOG: Eastern cooperative oncology group
cDNA: complementary DNA
qRT-PCR: quantitative Real Time PCR
GAPDH: glyceraldehyde-3-phosphate dehydrogenase
SD: standard deviation
ROC: receiver operating characteristics curve
AUC ROC: area under the ROC curve
AUCRP: Area Under Precision-Recall Curve
RMSE: Root Mean Square Error
EMVI: extramural vascular invasion
RFMDA: Random Forest Mean Decrease in Accuracy
DFS: disease-free survival
anti-PD-1 inhibitor: programmed death 1 inhibitor
MSI: microsatellite instability
dMMR: mismatch repair–deficient

