## Supplementary material for "Exploring novel inflammation-related genetic and hematological predictors of response to neoadjuvant chemoradiotherapy in locally advanced rectal cancer": Supp. Table 2

**Supplementary Table 2. Performance of the composite scores with respect to various metrics.**

| **Variable** | **AUC^1^** | **AUCPR^2^** | **RMSE^3^** | **RFMDA^4^** |
| --- | --- | --- | --- | --- |
| Absolute basophil count | 0.688 | 0.725 | 0.678 | 0.006 |
| Absolute eosinophil count | 0.669 | 0.705 | 0.574 | 0.003 |
| Absolute monocyte count | 0.686 | 0.691 | 0.485 | 0.002 |
| Neutrophil-to-monocyte ratio | 0.688 | 0.688 | 0.627 | 0.020 |
| ***Score 1*** |  |  |  |  |
| Absolute basophil count | 0.706 | 0.720 | 0.539 | 0.013 |
| Absolute eosinophil count |  |  |  |  |
| ***Score 2*** |  |  |  |  |
| Absolute basophil count | 0.720 | 0.731 | 0.486 | 0.011 |
| Absolute monocyte count |  |  |  |  |
| ***Score 3*** |  |  |  |  |
| Absolute eosinophil count | 0.711 | 0.754 | 0.555 | 0.012 |
| Absolute monocyte count |  |  |  |  |
| ***Score 4*** |  |  |  |  |
| Absolute basophil count | 0.724 | 0.760 | 0.585 | 0.009 |
| Absolute eosinophil count |  |  |  |  |
| Absolute monocyte count |  |  |  |  |
| ***Score 5*** |  |  |  |  |
| Neutrophil-to-monocyte ratio | 0.427 | 0.504 | 4.507 | 0.0001 |
| Absolute monocyte count |  |  |  |  |
| ***Score 6*** |  |  |  |  |
| Neutrophil-to-monocyte ratio | 0.580 | 0.634 | 1.909 | 0.003 |
| Absolute eosinophil count |  |  |  |  |
| ***Score 7*** |  |  |  |  |
| Neutrophil-to-monocyte ratio | 0.647 | 0.667 | 0.766 | -0.001 |
| Absolute basophil count |  |  |  |  |
| ***Score 8*** |  |  |  |  |
| Neutrophil-to-monocyte ratio | 0.485 | 0.576 | 6.382 | 0.004 |
| Absolute monocyte count |  |  |  |  |
| Absolute eosinophil count |  |  |  |  |
| ***Score 9*** |  |  |  |  |
| Neutrophil-to-monocyte ratio | 0.461 | 0.544 | 4.978 | 0.002 |
| Absolute monocyte count |  |  |  |  |
| Absolute basophil count |  |  |  |  |
| ***Score 10*** |  |  |  |  |
| Neutrophil-to-monocyte ratio | 0.616 | 0.669 | 2.402 | 0.006 |
| Absolute eosinophil count |  |  |  |  |
| Absolute basophil count |  |  |  |  |
| ***Score 11*** |  |  |  |  |
| Neutrophil-to-monocyte ratio | 0.515 | 0.598 | 6.875 | 0.006 |
| Absolute monocyte count |  |  |  |  |
| Absolute eosinophil count |  |  |  |  |
| Absolute basophil count |  |  |  |  |

*^1^Area Under Curve; ^2^Area Under Precision-Recall Curve; ^3^Root Mean Square Error; ^4^Random Forest Mean Decrease in Accuracy;*
