## Supplementary material for "Exploring novel inflammation-related genetic and hematological predictors of response to neoadjuvant chemoradiotherapy in locally advanced rectal cancer": Supp. Table 1

**Supplementary Table 1. Comparison of genes expression of patients with pCR and those with TRG4 postoperative category.**

| **Characteristic** | **The response to treatment** | | |
| --- | --- | --- | --- |
|  | **TRG1** | **TRG4** | **Wilcoxon rank sum test** |
| ***IL6*** |  |  |  |
| N (%) | 8/13 (61.5%) | 9/10 (90.0%) |  |
| Mean (SD) | 9.5 (13.5) | 1.0 (0.9) | *ns* |
| Median (Range) | 3.2 (0.03-36.1) | 0.6 (0.05-2.1) |  |
| ***CYBB*** |  |  |  |
| N (%) | 9/13 (69.2%) | 9/10 (90.0%) |  |
| Mean (SD) | 3.3 (3.9) | 1.1 (1.1) | *ns* |
| Median (Range) | 1.6 (0.6-10.3) | 0.8 (0.1-3.5) |  |
| ***CXCL9*** |  |  |  |
| N (%) | 9/13 (69.2%) | 9/10 (90.0%) |  |
| Mean (SD) | 1.4 (0.7) | 0.9 (0.6) | *ns* |
| Median (Range) | 1.6 (0.5-2.4) | 1.0 (0.1-1.9) |  |
| ***IDO1*** |  |  |  |
| N (%) | 9/13 (69.2%) | 8/10 (80.0%) |  |
| Mean (SD) | 1.7 (1.0) | 0.8 (0.4) | *p*=0.036 |
| Median (Range) | 1.7 (0.6-3.6) | 0.7 (0.2-1.6) |  |
| *Total* | 13 (100%) | 10 (100%) | *-* |

*ns - not statistically significant*
